# Mental Health Promotion in Victims of Colombian Armed conflict: a Scoping Review

**DOI:** 10.1101/2024.10.17.24315704

**Authors:** Lina María González Ballesteros, Luis Eduardo Mojica, Jennifer Clavijo Marín, Andrés Felipe Ortiz, Mariana Vásquez Ponce, Sofía Pérez-Lalinde, Nicolas León Sanabria, Luisa María Carrera Silva, Andrea Clavijo Álvarez

## Abstract

**Objective:** To explore mental health promotion interventions for victims of Colombian armed conflict.

**Design:** Scoping review.

**Methods:** Searches were conducted using indexed and free terms regarding mental health promotion interventions for victims of Colombian armed conflict in different databases, including MEDLINE, EMBASE, Scielo, Cochrane Central Register of Controlled Trials, LiLACS, Google Scholar, and PsycArticles. We included primary and secondary research sources, including clinical trials, randomized controlled trials, qualitative research, online journals, meta-analyses, scoping reviews, and systematic reviews, as well as government reports and articles about implementing mental health promotion strategies for victims of Colombian armed conflict.

**Results:** Our search returned 317 identified articles through database exploration. Four more articles were included through literature research and another four records were included through gray literature and government reports. Retrieved citations were screened using Rayyan QCRI. A total of 259 records were left after duplicate removal. Members of the group then performed a paired-blind screening, which left us with 16 articles to be included in the analysis. Data analysis was conducted by authors in pairs using custom tools developed by the research team. After reviewing the articles, only six met the inclusion criteria established for this scoping review. Among these, two of the interventions addressed were randomized clinical trials that demonstrated effects on depression, anxiety, and post-traumatic stress disorder (PTSD). Additionally, one of these trials assessed impairments in functionality.

**Conclusion:** A total of six articles demonstrated the effectiveness of mental health promotion and primary prevention interventions in Colombian victims of armed conflict. However, only two of the studies were randomized clinical trials, and methodological limitations were evident, including loss in sample follow-up, challenges in comparing populations, and use of non-standardized scales across different studies. This hinders the possibility of a unified assessment of these interventions and highlights the need for bigger study samples and use of tools that facilitate access to programs and one-year follow-ups to enhance interventions government organizations employ to address these mental health needs.

## INTRODUCTION

Recognition of the importance of mental health has grown significantly in recent years, given its impact on people’s general well-being and as an indicator of a country’s development. The World Health Organization (WHO) describes ‘mental health’ as a “state of individual, community, and socio-economic mental well-being that enables people to cope with the stresses of life, realize their abilities, learn well and work well, and contribute to their community” (WHO, 2022). Furthermore, it describes that ‘mental health promotion’ encompasses actions aimed at modifying social determinants to impact individual and collective health positively (WHO, 1986). Understanding these concepts is essential since, although inequity in terms of disability and mortality in people with mental disorders has been documented and prevention strategies have been created to prevent these disorders, there is a lack of information available on negative mental health outcomes and promotion actions in this area (WHO, 2021).

Worldwide, exposure to armed conflict increases vulnerability to psychosocial distress and mental disorders, with victims experiencing up to 20% higher prevalence of mental disorders than the general population (Charlson et al., 2019). Depression, anxiety, and PTSD are among the most common disorders, and psychosomatic disorders resulting from physical impairment are also documented (Piñeros et al., 2021)), along with an increased risk of suicide and alcoholism (Restrepo & Padilla-Medina, 2023; Trujillo et al., 2021). However, pathologies are not the only negative outcomes in this population; the uprooting derived from displacement and the death of family members inevitably leads to personal, cultural, and collective identity crises, rupture of social ties, and limitations to establishing new support networks in resettled territories, all of which perpetuate community isolation, social dysfunction, and deterioration of mental health (Comisión de la verdad, 2022).

With this background, mental health promotion has become a necessity for people affected by armed conflict. Strategies for preventing revictimization worldwide include peer support groups, psychoeducation, and community intervention (Tol et al., 2013). Coldiron et al. also describes how brief psychotherapeutic interventions offered by Médecins Sans Frontières in populations in China, Colombia, and Palestine have resulted in significant improvement in the reduction of affective symptoms, concluding that, even during conflict, it is possible to provide high quality mental health care. An analysis of the short and medium-term consequences of Operation Cast Lead on the population of the Gaza Strip, from 2007 to 2011, showed the importance of carrying out a constant evaluation of affected people in order to make appropriate adjustments to mental health care programs in contexts of chronic violence (Llosa et al., 2012).

In Colombia, armed conflict has spanned at least two centuries, characterized by civil wars, bipartisan struggles, and violence fueled by political movements. Despite attempts to seek an end to the violence, armed conflict in Colombia has had a significant historical impact on the country with international ramifications (Molando Bravo, 2015). The armed conflict has been characterized by direct and constant harmful effects on the civilian population, with multiple negative economic, cultural, and health results. The Unit for the Attention and Integral Reparation of Victims (UARIV) reported 9,555,446 victims of Colombian armed conflict (Víctimas, 2023). However, there is evidence of underreporting, especially in minority populations such as indigenous, Afro-Colombian, Palenquero, Raizal, and Romani populations. Among the victims, it has been documented that 5.47% had psychosocial disabilities (Social, 2020; Víctimas, 2023),. As of June 2021, the Internal Displacement Observatory (IDMC) estimated that approximately 5,235,064 people had been internally displaced by the armed conflict (Cazabat & O’Connor, 2021), making Colombia the country with the second highest number of victims of internal displacement in the world at the time.

The effects of violence in the country do not differ from those reported in the world. Victims have a 74% higher chance of suffering from a mental disorder compared to non-victims. Among socioeconomic groups, low-income populations experience a disproportionately negative effect and an inequality in the presentation of and attention to mental disorders (Cuartas et al., 2019; Ministerio de Salud, 2015). An exploratory analysis by S. Leon-Giraldo; et al. evaluated data collected in the 2014 Conflict, Peace and Health Survey (CONPAS) conducted in the Meta region (among the most affected by internal displacement) and the 2015 National Mental Health Survey (ENSM), and found that people who self-reported as internally displaced presented a higher probability of having a mental health disorder (14%-21.8%) compared to the general population. Suicidal ideation at the national level had a higher probability of being presented by 11.9% (95% CI, 9.3-14.1), while at the regional level there was 21.4% higher probability of reported anxiety (95% CI, 9.3-14.1) and a 12.4% higher probability of reported depression (95% CI, 9.5-15.7), compared to lower levels at the national level. This data does not differ from studies carried out in other Colombian regions. Londoño A, et al., for instance, in a cross-sectional study comparing two populations, described an increased prevalence of mental health disorders in a population directly exposed to violent events compared with a non-exposed population. Additionally, it has been estimated that up to 11% of forcibly displaced women are victims of rape, and they experience domestic violence at roughly twice the rate of displaced men (Tol et al., 2013)((Agudelo-Vélez, 2018). Studies have also shown that in contexts of conflict, the mental health of caregivers is compromised, limiting their ability to support and raise children effectively, which heightens the risk of alterations in the psychological development of children (Ballesteros et al., 2021).

In light of the profound effects of violence on the Colombian population, the Psychosocial and Integral Health Program for Victims (PAPSIVI) was implemented. PAPSIVI stands as a comprehensive framework of activities, procedures, and interdisciplinary interventions, designed and championed by the Ministry of Health and Social Protection. Understanding the significance of PAPSIVI is crucial, as it embodies a government-backed mechanism for addressing direct psychosocial and health consequences and serves as a model for holistic care (Colombia Government, 2017). Evaluating PAPSIVI’s effectiveness in this capacity is particularly relevant, as it sheds light on how structured governmental programs can influence mental health outcomes in post-conflict settings.

In Colombia, reported interventions in mental health promotion are scarce. Main publications on the subject include qualitative studies conducted in regions affected by armed conflict, which have shown the importance of community social support in the preservation of mental health. In the Médecins Sans Frontières study, it was highlighted that interventions must be adapted to context, integrating culturally-sensitive concepts of suffering, treatment, and illness (Sanchez-Padilla et al., 2009).

As it has been established, to this date there is a clear insufficiency in literature on mental health promotion interventions for victims of armed conflict in Colombia, as well as a lack of clarity on the effectiveness of the few interventions that have been applied. Therefore, the purpose of this scoping review is to fill this research gap and expand the understanding of impactful and effective interventions for mental health promotion among Colombian armed conflict survivors.

## METHODOLOGY

### 1. Search strategy

A scoping review was conducted using the methodological framework described by the Joanna Briggs Institute and the Preferred Reporting Items for Systematic Reviews and Meta-Analyses extension for scoping reviews (JBI, n.d.). The basis for the literature search was the following research question: Taking into account the unique context, problems, and challenges faced by this population, what is the scope, extent, and nature of the research evidence on the effectiveness of mental health promotion interventions among victims of Colombian armed conflict?

Relevant MeSH, EMTREE, and free terms were selected based on this research question. Searches were performed in the MEDLINE (PubMed), EMBASE (EMBASE), Scielo, Cochrane Central Register of Controlled Trials (Ovid), LiLACS (BVS), and PsycArticles (PsycNet) databases. Gray literature was reviewed in ProQuest Dissertations and Theses Global (ProQuest). In addition, government reports and articles related to the implementation and effectiveness of PAPSIVI psychosocial strategies and those of other authors were searched and obtained from a Google Scholar search (Table 1).

**Table 1.** Results of literature search on the database used.

| Literature Search with Keywords | Data Base | Results |
| --- | --- | --- |
| (Armed conflict OR warfare OR refugee*) AND (Victim*) AND Colombia OR Colombian*) AND (Mental health) ) AND (Armed conflict OR warfare OR refugee*) AND (Victim*) AND Colombia OR Colombian*) AND (Mental health)) | Pubmed (Medline) | 26 |
| Conflicto armado) AND (Colombia) AND ("Salud mental") | Scielo | 23 |
| Conflicto armado) AND (Estrategias salud mental) AND Colombia) | Scielo | 5 |
| 'armed conflict'/exp OR 'armed conflict') AND ('colombia'/exp OR colombia) AND 'mental health' | EMBASE | 85 |
| 'armed conflict'/exp OR 'armed conflict') AND 'victims' AND colombia*' AND ('mental health promotion'/exp OR 'mental health promotion' OR 'mental health intervention*' OR 'mental health strategy*') | EMBASE | 4 |
| Armed conflict) AND (Colombia) AND (mental health) | PsycNet<br>(PsycArticles) | 61 |
| Armed conflict OR warfare OR refugee*) AND (Victim*) AND Colombia OR Colombian*) AND (Mental health) | Cochrane Central<br>Register of | 4 |
|  | Controlled Trials<br>(Ovid) |  |
| CONFLICTO ARMADO" AND "COLOMBIA" AND "SALUD<br>MENTAL" AND ( db:("LILACS") AND la:("es" OR "en")) | LiLACS (BVS) | 41 |
| victims colombian armed conflict) AND (mental health<br>promotion) AND effectiveness AND context-specific problems | ProQuest<br>Dissertations and<br>Theses Global<br>(gray literature) | 68 |
| PAPSIVI" AND "effectiveness" OR "evaluation" | Google scholar<br>(gray literature) | 6 |

The presentation of the data found, reviewed, and analyzed is presented in a Preferred Reporting Items for Systematic Reviews and Meta-Analysis extension for scoping review (PRISMA-ScR) flow diagram (Figure 1).

**Figure 1.**
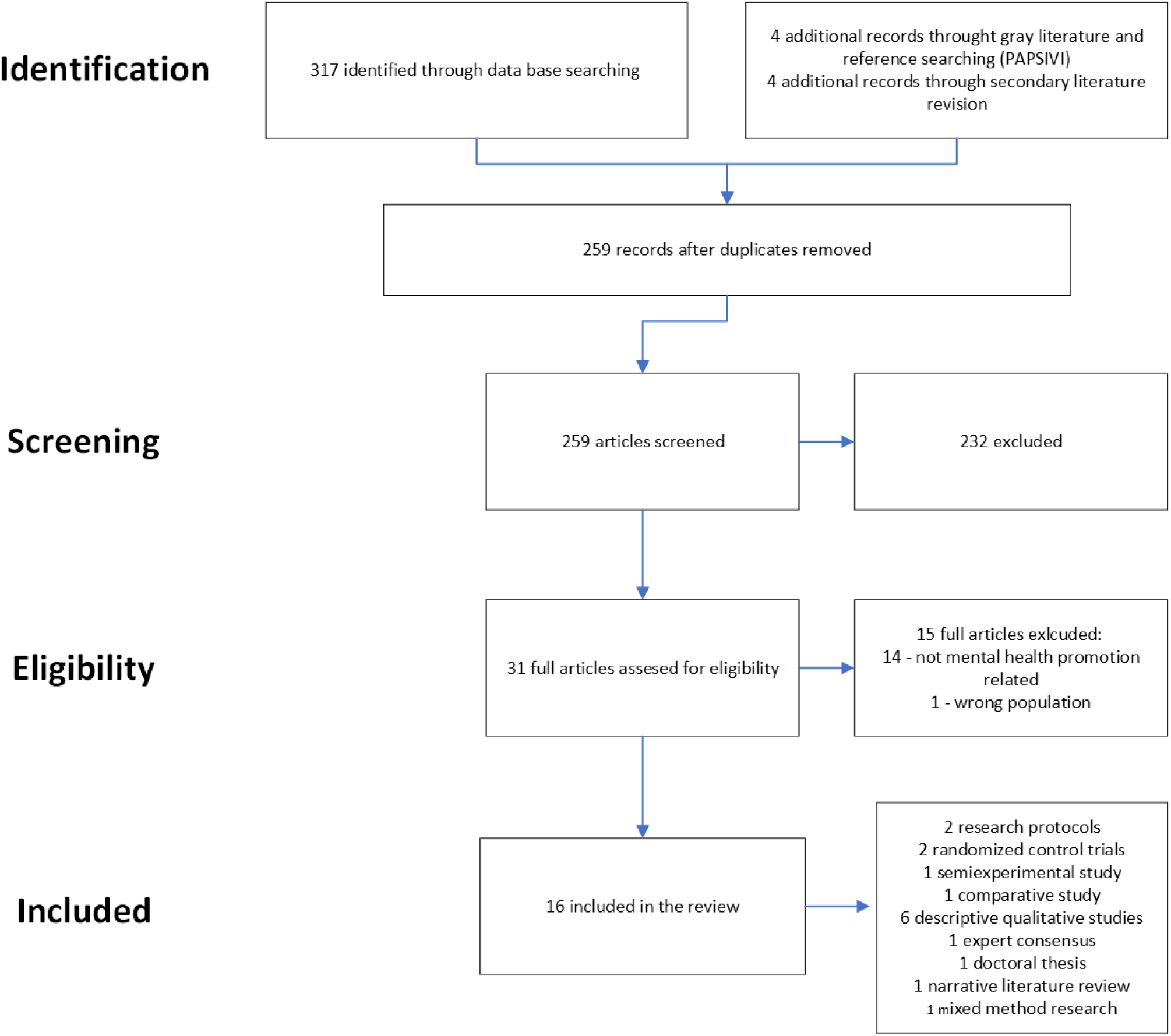
PRISMA diagram.

### 2. Eligibility Criteria

Once the literature search was completed, all citations retrieved were uploaded to Rayyan QCRI, a tool used to automatically eliminate duplicates. Subsequently, initial screening was performed based on examination of the title and abstract of each article by two reviewers, with the inclusion of a third in the case of disagreement. Articles approved by two reviewers were included. Within the inclusion criteria, original research articles specific to mental health promotion interventions in victims of armed conflict in Colombia were considered, including, but not limited to, clinical trials and other primary, secondary, and tertiary sources, such as governmental reports.

### 3. Charting, Collection, Summarizing, and Analyzing data

After the initial screening, the full texts were read and summarized into a database of tables with headings for: title, objective of the article, population, concept, context, type of evidence, country, participants in the case of a clinical trial, and details of the results. With these tables, a new review was carried out in which the researchers confirmed whether or not the inclusion criteria of the articles reviewed was met.

A new database was created with the documents that met this criteria, including, in addition to the data previously described, details of the intervention carried out, evaluation tools used, and description and validity of the intervention, as well as details of the results and points to be highlighted by each researcher on the article.

To develop the analysis, discussion, and conclusions, all authors read the final articles. Researchers included a specialist in psychiatry, three psychiatric residents and five medical students, with the aim of achieving a diversity of perspectives. Subsequently, two virtual meetings were held, which were recorded and transcribed. The first phase of the review involved a rigorous assessment of the eligibility of the articles for inclusion. This evaluation considered the following factors: the context of the interventions, the methodological approaches employed, the presented results, and any existing consensus discussions. The selected studies were then discussed in detail in a second meeting, leading to final conclusions reached through a collaborative consensus process.

## RESULTS

Our search yielded a pool of 317 articles through exploration of databases, with an additional four records sourced from gray literature and government reports contributing to the diversity of the dataset, and four more articles were included through literature research. After using Rayyan QCRI to remove duplicates, 259 records were screened through a rigorous paired-blind screening process undertaken by the research team, resulting in the detailed assessment of 30 articles for eligibility, of which 15 were excluded because they did not meet inclusion criteria.

Ultimately, 16 articles were selected for the analysis phase including two research protocols, two randomized control trials, one mixed-method research study, one quasi-experimental study, six descriptive qualitative studies, one expert consensus, one doctoral thesis, one narrative literature review, and one observational evaluation study. Out of the reviewed studies, the research team identified ten that did not meet the inclusion criteria. (Table 2).

**Table. 2.** Excluded articles.

| Number | References | Title | Country | Year | Setting | Study type | Study focus |
| --- | --- | --- | --- | --- | --- | --- | --- |
| 1 | Sarmiento-Marulanda et al. | Psychosocial rehabilitation experiences of women victims of armed conflict in Montes de María, Colombia | Colombia | 2021 | Montes de María region | Qualitative study with a phenomenological approach | Perception and experiences of women victims of the colombian armed conflict regarding psychosocial rehabilitation of the Victims Law |
| 2 | Giebel et al. | Understanding post-conflict mental health needs and co-producing a community-based mental health intervention for older adults in Colombia: a research protocol | Colombia | 2022 | Turbo, Antioquia | Mixed method research | Understanding the effect of stressful life events on the mental health of elders and how its impact can be reduced by a co-produced community intevrention |
| 3 | Bonilla-Escobar et al. | Do not forget culture when implementing mental health interventions for violence survivors | Colombia | 2017 | Pacific region | Qualitative reasearch | Understand how afrocolombian culture aids communities to cope with the effects of the colombian armed conflict and how it can be implemented when applying mental health interventions on this groups |
| 4 | Bonilla-Escobar et al. | Evaluation of Two Community-based Mental Health Interventions for Violence-Displaced Afro-Descendants in Colombia | Colombia | 2021 | Pacific region | Follow up study to randomized control trial | Measure the effects of the CETA and NCGT interventions in reducing PTSD, depression, anxiety and level of function impairment |
| 5 | Kalksma-Van Lith | Psychosocial interventions for children in war-affected areas: The state of the art. | Colombia, Lebanon, Mexico, China, Palestine and Russia. | 2007 | War affected areas | Review of qualitative and quantitative research | Review the literature available on psychosocial assitance to chlidren in war affected areas |
| 6 | Rodríguez, De la Torre, Miranda | Mental health in situations of armed conflict (La salud mental en situaciones de conflicto armado) | Latin American countries | 2002 | Countries in Latin America in situations of armed conflict | Review of literature and epidemiological data | Describe the psychosocial consequences of armed conflict and violence, the epidemiological data regarding psychiatric disorders resulted from trauma, and analyze the lines of action in mental health plan focused on the youth |
| 7 | Pacichana-Quinayáz SG, Osorio-Cuéllar GV, Bonilla-Escobar FJ, Fandiño-Losada A, Gutiérrez-Martínez MI | Common Elements Treatment Approach based on a Cognitive Behavioral Intervention: implementation in the Colombian Pacific | Colombia | 2023 | Buenaventura and Quibdó, Colombia. | Qualitative through individual in-depth interviews with LPCWs | Describe the implementation of CETA for Afro-Colombian victims of violence, based on perceptions of the therapy implementers (LPCWs) in Buenaventura and Quibdó, Colombia. |
| 8 | Ametaj, Amantia | Assessing contextual factors for implementing evidence-based care in a middle income country in the wake of conflict: The Colombian mental health system | Colombia | 2019 | Colombia | Qualitative reasearch | Asses the capacity of the Colombian health system to implement evidence-based psychological mental health care services for victims of the armed conflict |
| 9 | Sera, Daniel Ruiz; Marchand, Ines | Agape: A reconciliation initiative by members of civil society and former child-soldiers. | Colombia and Canada | 2011 | Colombia | Qualitative reasearch | To provide a space for reconciliation among armed conflict victims; promote training oportunities for children-soldiers in an area of their interest; break the paattern of rejection that often accompanies the youngsters; and to offer the child-soldiers an opportunity to experience life in a context of peace. |
| 10 | Osorio-Cuellar GV, Pacichana-Quinayáz SG, Bonilla-Escobar FJ, Fandiño-Losada A, Gutiérrez-Martínez MI | Perceptions about implementation of a Narrative Community-based Group Therapy for Afro-Colombians victims of Violence | Colombia | 2017 | Buenaventura and Quibdó, Colombia. | Qualitative research | To describe the process of implementation and results of the NCGT intervention for victims of violence, in the municipalities of Buenaventura and Quibdó - Colombia, from the perspective of LPCW (Lay Psychosocial Community Workers) and their supervisors |

All six of the qualitative studies were ultimately excluded. One study was excluded because it did not specifically focus on interventions (Bonilla-Escobar et al., 2017). Another study was excluded because it did not include any mental health interventions (Sarmiento-Marulanda et al., 2021). Two were excluded because they were part of the same intervention as a third that ultimately was selected (Osorio-Cuellar et al., 2017; Pacichana-Quinayáz et al., 2016). Another article was excluded because the study focused on the barriers to accessing the health system rather than the effectiveness of a specific mental health promotion strategy (Ametaj, 2019). Finally, a qualitative study was excluded because it did not present an evaluation of the intervention effectiveness (Ruiz-Serna & Marchand, 2011).

The narrative literature analysis was ultimately excluded because it lacked a specific focus on interventions (Kalksma-Van Lith, 2007). The expert consensus was excluded because it didn’t present a direct intervention (Rodríguez et al., 2002). Finally, two sources were excluded because they were research protocols (Bonilla Escobar et al., 2018; Giebel et al., 2022).

Six articles ultimately met the inclusion criteria for review. These included two randomized controlled trials (RCTs), one quasi-experiment, one comparative study, one doctoral thesis, and one mixed-method research study (Table 3).

**Table 3.**
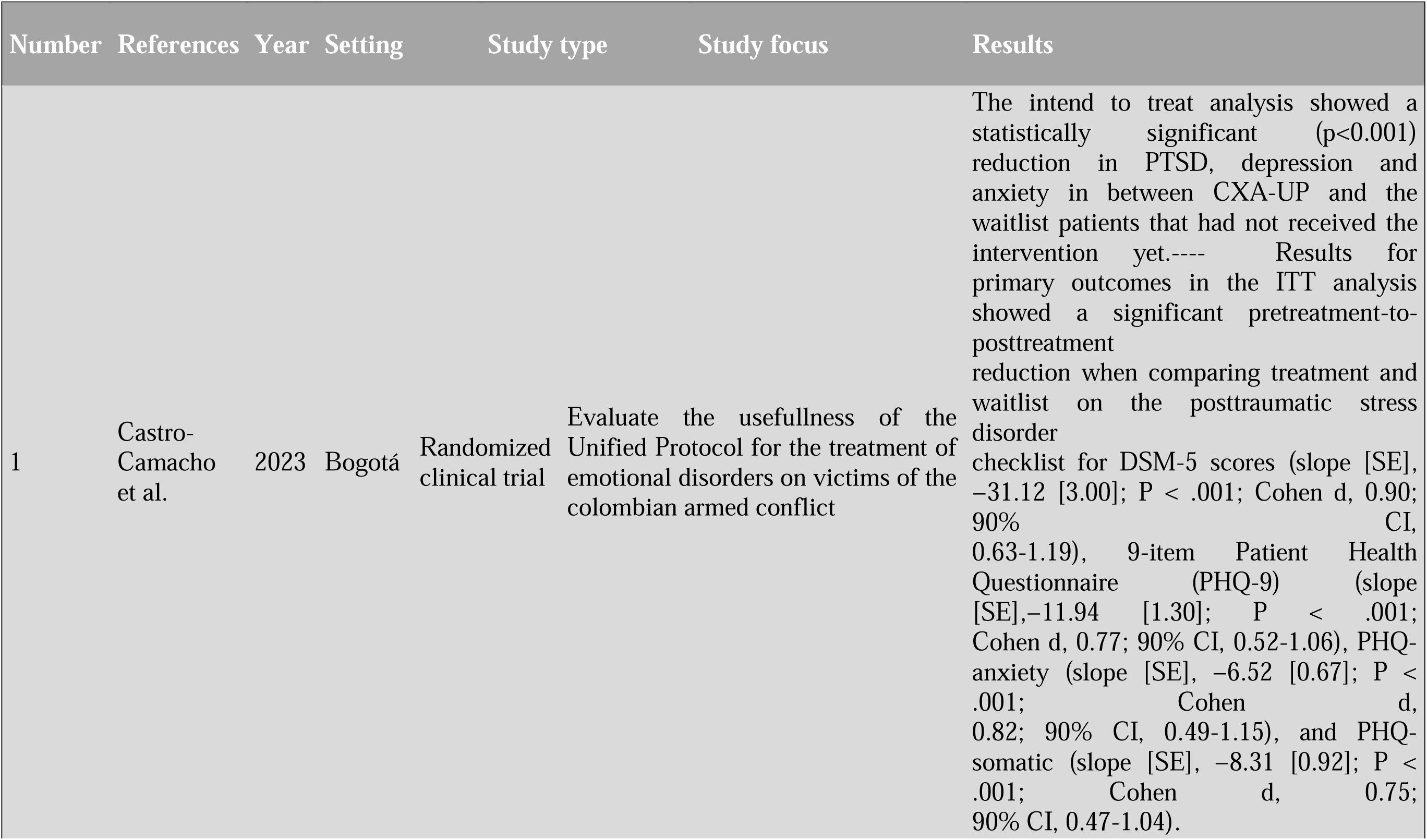
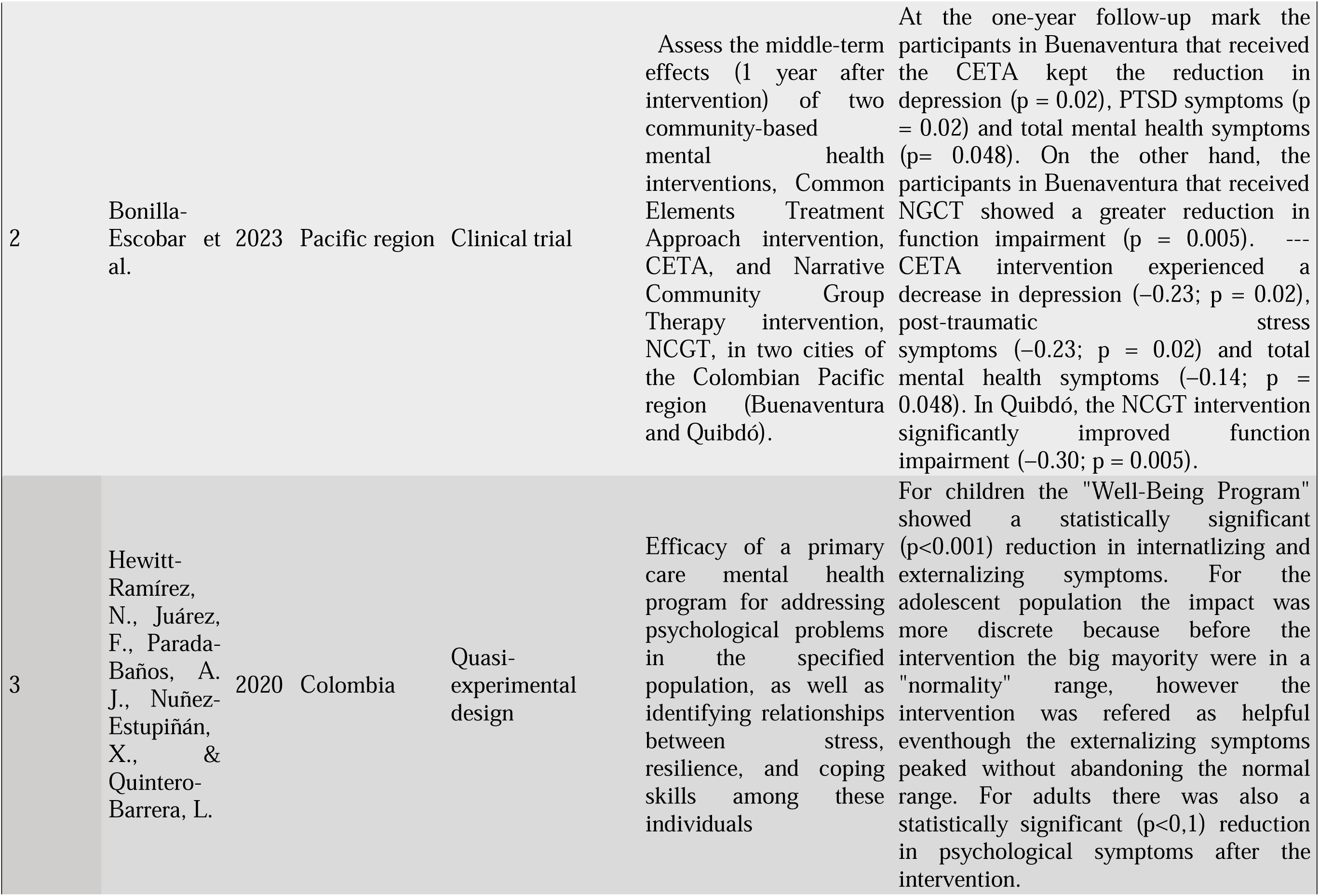
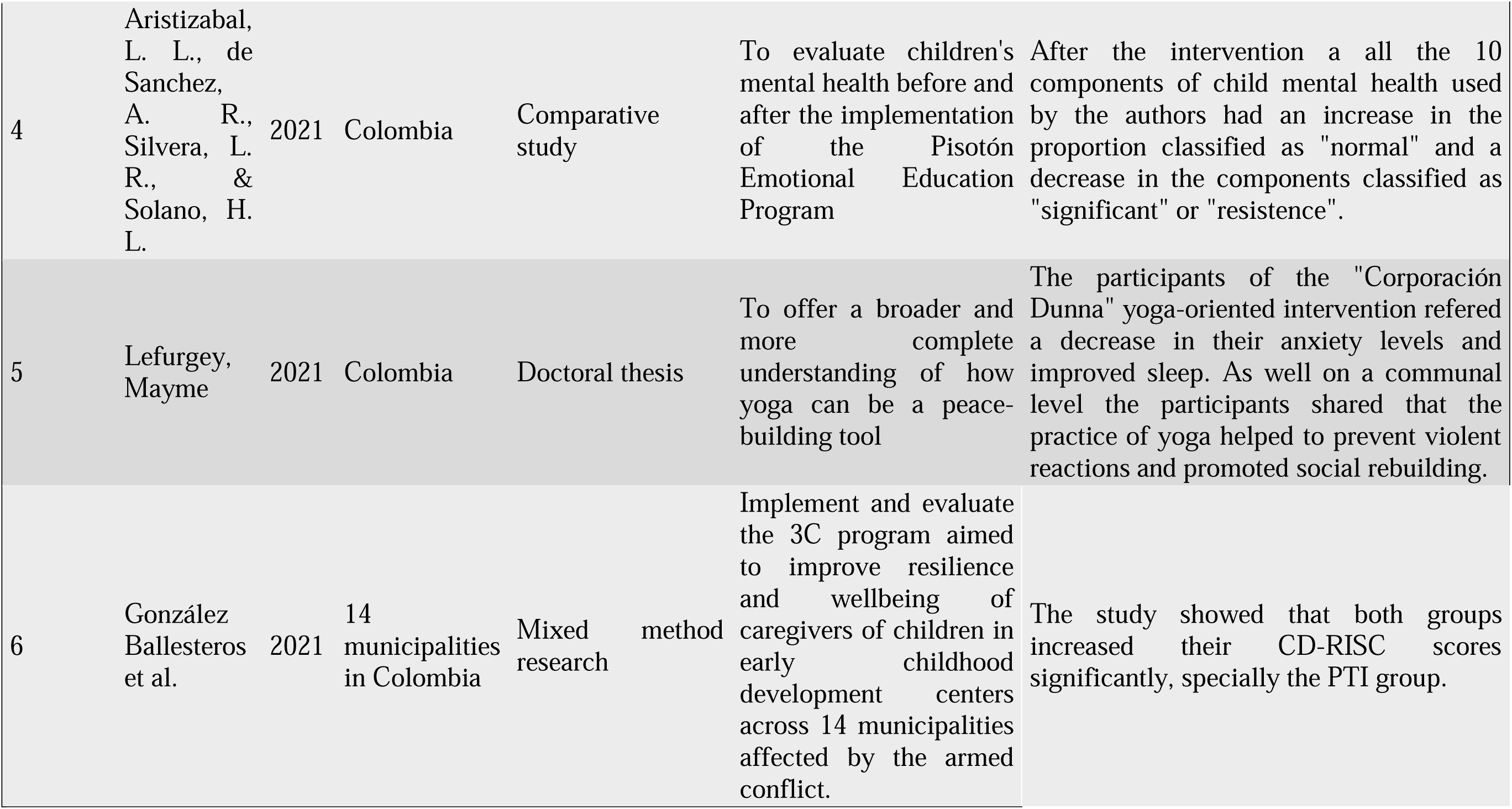
Articles included.

One of the included RCTs, “Effects of a Contextual Adaptation of the Unified Protocol in Multiple Emotional Disorders in Individuals Exposed to Armed Conflict in Colombia: A Randomized Clinical Trial” (Castro-Camacho et al., 2023) demonstrated the statistical effectiveness of the Contextual Adaptation of the Unified Protocol (CXA-UP) in reducing PTSD (Cohen d, 0.90; CI, 0.63-1.19), depression symptoms (Cohen d, 0.77; CI, 0.52–1.06), anxiety (Cohen d, 0.82; CI 0.49–1.15), and psychosomatic symptoms (Cohen d, 0.75; CI 0.47-1.04), while enhancing overall functioning, as assessed through primary and secondary scales. Due to a significant participant sample loss for follow-up, impacting the intent-to-treat and per-protocol analyses, the latter showed a larger effect, but all observed effects were considered substantial. Importantly, the positive impact was sustained at the 3-month follow-up assessment.

The second RCT, “One-Year Outcomes of Community-Based Mental Health Interventions for Afro-Colombian Survivors of Armed Conflict and Displacement” (Bonilla Escobar et al., 2023) is the latest publication in the ACOPLE study series. It compares the efficacy of the Common Elements Treatment Approach (CETA), a cognitive-behavioral therapy approach, with Narrative Community Group Therapy (NGCT), based on oral narration and group cooperation, assessing PTSD, depression, anxiety, and functional impairment in two Pacific Colombian cities – Buenaventura and Quibdó – one year post-intervention. The study series used the Hopkins Symptom Checklist and Harvard Trauma Questionnaire as instruments for the outcome assessment before the intervention, two-weeks after completion, and one-year following. It found positive effects for both interventions. Notably, while an increase in symptoms occurred between the 2-week follow-up and one-year assessment, it did not negate overall improvement from baseline scores. CETA showed reductions in PTSD (–0.23; P=0.02) and depression symptoms (– 0.23; P= 0.02), along with improved function, whereas NGCT enhanced overall function (–0.30; p= 0.005) and was particularly effective for depression symptoms in Buenaventura. Authors attribute result diversity to city-specific violence structures, altering conflict exposure, and varying levels of insecurity and poverty. However, limitations include unaccounted for psychosocial care or treatment, which impacted result interpretation, and a small sample size especially given a 43.8% participant loss between the initial intervention and the one-year follow-up.

The quasi-experiment titled “Efficacy of a Primary Care Mental Health Program for Victims of Armed Conflict in Colombia” (Hewitt-Ramírez et al., 2020) evaluated the 21-month-long Well-Being Program, which included community establishment, mental health care facilities, therapy, psychosocial support, promotion, prevention activities, and a needs analysis. The intervention zone targeted individual, family, and community levels, offering clinical and psychosocial support through nine, 2-hour educational sessions, with the aim of promoting healthy lifestyles and enhancing social competencies and life skills in children, adolescents, teachers, and student families across four schools. Finally, it investigates the program’s impact on reducing psychological problems and explores the relationship between stress, resilience, and coping skills, through various scales for each age group. Key findings reveal a significant decrease in psychological issues and an improvement in participants’ coping mechanisms.

The comparative study, *“Pisotón, Emotional Education Program for the Promotion of Children’s Mental Health in Territories Affected by Colombian Armed Conflict: A Comparative Study”* (Aristizabal et al., 2021)was conducted to assess the effects of an emotional education program for children. The program consisted of 11 modules, each employing four different techniques (storytelling, psychodrama, home play, and experiential storytelling). While improvements were observed in areas like handling complex emotions, independence, coping mechanisms, and family relationships, the study recognized some methodological limitations, like the use of the Duess Fables Test, an instrument used to identify potential crises or developmental conflicts, which lacks validation in Colombia.

The doctoral thesis, ***“Yoga as Embodied Peacebuilding: Moving through Personal, Interpersonal, and Collective Trauma in Post-conflict Colombia”*** (Lefurgey, 2021)explores the author’s engagement with the non-profit organization “Corporación Dunna,” employing integral yoga for bottom-up peacebuilding in violence-affected communities. Through a transrational, relational, and embodied approach, the author conducted 75 semi-structured interviews over three months. The diverse interviewees included citizens selected through snowball sampling, incarcerated youth, Dunna corporation staff, and experts. The study revealed individual benefits like reduced anxiety and improved sleep, while at familial and communal levels, yoga practices were credited with preventing violent reactions and fostering social rebuilding.

Finally, the mixed-method research study, titled “***Evaluating the 3Cs Program for Caregivers of Young Children Affected by Armed Conflict in Colombia”*** (Ballesteros et al., 2021)investigated the impact of the Contigo, Conmigo, Con Todos (3Cs) program on self-reported resilience among caregivers of young children in conflict-affected areas. This mental health intervention consisted of two modules: a skill-building program (SBP) and a psychotherapy intervention (PTI). The research aimed to assess the influence of program attendance on resilience, as measured by the CD-RISC score. Specifically, the PTI group, which had a lower baseline resilience score demonstrated a significant improvement. Additionally, a higher-than-average attendance in SBP was associated with a statistically-significant increase in CD-RISC scores.

## DISCUSSION

This study aimed to identify the scope, extent, and nature of existing evidence on the effectiveness of mental health promotion interventions among victims of Colombian armed conflict. Consistent with existing international literature, findings show that there is a high rate of mental disorders in Colombian populations exposed to armed conflict, primarily anxiety disorders, depression, and post-traumatic stress disorder (Cuartas et al., 2019).

The articles included in this review aimed to assess the effectiveness of mental health interventions, including group cognitive behavioral therapy, narrative community group therapy, and psychodynamic approaches, over different outcomes in victims of armed conflict. Each article evaluated a different type of intervention and measured results with different assessment tools. Positive results were found with regards to combatting some of the most common mental health disorders such as PTSD, anxiety, and depression throughout two of the articles (Bonilla Escobar et al., 2023; Hewitt-Ramírez et al., 2020).

However, these results should be interpreted with caution due to a lack of evidence, as most of them came from non-randomized research with various limitations, the most common being a high rate of participant loss, which led to smaller follow-up samples, as well as a lack of standardized and validated scales.

Considering the limitations previously mentioned, it is imperative to adopt diverse strategies for future research interventions. These should include the use of country-specific, validated tools and strategies to reduce participant dropout, such as enhancing transportation, providing flexible scheduling, and maintaining continuous communication with participants. Additionally, it’s crucial to explore various intervention methods, particularly ones that leverage technology and virtual platforms. This approach should increase access for individuals in remote or hard-to-reach areas and tailor interventions to meet unique cultural needs and address potential trauma triggers common in these populations. Enhanced funding and resources are essential for these improvements, highlighting the critical role of government and international agencies in supporting the mental health of armed conflict victims.

Finally, long-term follow-up, extending to at least one year, is necessary to assess the enduring impact of these interventions on individuals and communities. This underscores the imperative for more comprehensive and robust research to fully comprehend and augment the effectiveness of mental health interventions in conflict-impacted regions.

## LIMITATIONS

A notable limitation in our study is the subjective nature of identifying individuals or communities as ‘victims.’ While the World Society of Victimology defines a victim as “someone who has suffered harm through violations of criminal laws, including abuses of power,” the self-identification of a person or community as a ‘victim’ is influenced by sociocultural factors like social stigma and fear of labels. In our investigation, this poses a challenge, as individuals affected by armed conflict, despite being victims, may not readily identify as such, potentially impacting the outcomes presented in our review.

## CONCLUSION

A total of six articles demonstrated the effectiveness of mental health promotion and primary prevention interventions in Colombian victims of armed conflict. However, only two of the studies were randomized clinical trials, and methodological limitations were evident, including loss in sample follow-up, challenges in comparing between populations, and use of non-standardized scales across different studies. This hinders the possibility of a unified assessment of these interventions and highlights the need to use bigger study samples and employ tools that facilitate access to programs and one-year follow-ups in order to enhance the interventions that government organizations use to address these mental health needs.

## Data Availability

All data produced in the present study are available upon reasonable request to the authors
All data produced in the present work are contained in the manuscript
All data produced are available through an email to the authors

WHO: World Health Organization
UARIV: Unit for the Attention and Integral Reparation of Victims
IDMC: Internal Displacement Observatory
PTSD: post-traumatic stress disorder
CONPAS: Conflict, Peace and Health Survey
ENSM: National Mental Health Survey
PAPSIVI: Psychosocial and Integral Health Program for Victims
MPH: Mental Health Promotion
CXA-UP: Unified Protocol
CETA: Common Elements Treatment Approach
NGCT: Narrative Community Group Therapy

## DECLARATIONS

### Ethics Approval and Consent to Participate

The study received approval from the Psychiatry Department Research Committee from Pontifical Xavierian University

### Consent for Publication

Not applicable

### Availability of Data and Materials

Data is available upon reasonable request from the corresponding author.

## Acknowledgements

C. The authors would like to acknowledge Nubia Esperanza Bautista Bautista, Coordinadora del Grupo Convivencia Social y Ciudadanía, Dirección de Promoción y Prevención, and Ministerio de Salud y Protección Social.

